# Rare-Class Collapse in ECG-Based Ventricular Tachycardia and Fibrillation Detection: A Systematic Benchmark of Class-Imbalance Mitigation from Reweighting to Cascade Classification

**DOI:** 10.64898/2026.06.26.26356694

**Authors:** Kabi Raj Tiruwa

## Abstract

Ventricular tachycardia (VT) and ventricular fibrillation (VF) are the leading electrical causes of sudden cardiac death, but automated detection is limited by strong class imbalance, where lethal arrhythmias account for fewer than 22% of ECG segments. In this setting, standard classifiers can achieve high accuracy by predicting normal rhythm in most cases while missing many lethal events, a failure mode referred to as rare-class collapse. Six imbalance-handling approaches were evaluated: naive logistic regression, inverse-frequency reweighting, label-distribution-aware margin loss (LDAM), cost-sensitive training, two-stage cascade classification, and anomaly detection on 15,614 ECG segments from three PhysioNet databases (VTaC, VFDB, CUDB), with an overall normal-to-lethal ratio of 3.6:1. All methods were assessed at a fixed operating point of 95% specificity using recall, area under the precision–recall curve (AUPRC), and missed-lethal-event rate (MLER). The naive model achieved 45.1% recall (MLER = 0.549), missing 564 of 1,027 lethal events despite 84.1% accuracy. The two-stage cascade performed best, with 65.2% recall, AUPRC of 0.821, and MLER of 0.348, reducing missed events by 37% and achieving the highest decision-curve net benefit. Per-source analysis showed near-complete VF detection (recall up to 0.975) but much lower VT detection (recall 0.183), suggesting a feature-space limitation due to spectral similarity between organized VT and rapid sinus rhythm. Overall, the results show that evaluation metrics strongly influence the visibility of rare-class failure, and that cascade-based methods outperform simpler reweighting approaches for detecting lethal arrhythmias.

## 1 Introduction

Ventricular tachycardia (VT) and ventricular fibrillation (VF) are the main electrical causes of sudden cardiac death, and rapid intervention after rhythm onset is critical for survival [1]. Continuous electrocardiographic (ECG) monitoring in intensive care units, ambulatory Holter devices, and consumer wearables plays an important role in detecting these events early enough for timely treatment [1, 2]. A major challenge in automated VT/VF detection is that these life-threatening arrhythmias occur far less frequently than normal rhythms in ECG recordings, often several-fold in clinical monitoring datasets and by greater margins in population-level surveillance [3, 4]. This phenomenon, commonly termed the class imbalance problem [3, 4], can cause models to systematically under-detect minority-class events. In the context of lethal arrhythmia detection, this failure mode is referred to here as rare-class collapse to emphasize its clinical consequence: a missed VT/VF event is not a statistical error but a potentially fatal one. Conventional evaluation metrics can obscure this limitation. Accuracy is heavily influenced by the large number of normal rhythm samples, and the area under the receiver operating characteristic (ROC) curve may also appear favorable when negative samples greatly outnumber positive samples [5–7]. In contrast, precision–recall metrics and recall at a fixed clinical specificity provide a more meaningful assessment of performance because they focus on the minority class and the model’s ability to identify VT/VF events [5–7]. However, the most effective strategy for improving recall of these lethal arrhythmias under realistic class imbalance remains unclear.

A large body of research has advanced automated arrhythmia detection, ranging from traditional classifiers based on ECG morphology and interval features [8] to deep learning models such as convolutional and recurrent neural networks that have achieved cardiologist-level performance in beat and rhythm classification [9–12]. To address class imbalance, several approaches have been proposed, including synthetic oversampling [13], focal-loss and cost-sensitive networks [14–16], margin-based loss functions [17, 18], and two-stage or ensemble methods [19, 20]. Many of these approaches have reported strong performance on imbalanced ECG datasets [21, 22], including models developed specifically for shockable rhythm detection [23]. However, many studies evaluate models on rebalanced datasets or report metrics such as accuracy and F1 score rather than recall at a clinically relevant operating point. As a result, an important clinical measure—the number of lethal events missed at an acceptable false-alarm rate—is often not reported. In addition, different imbalance-handling methods are typically evaluated using different datasets, preprocessing strategies, and experimental settings, making direct comparison difficult. Consequently, it remains unclear which approach is most effective for detecting rare but life-threatening arrhythmias under realistic conditions.

Whether imbalance-handling methods generalise across ECG databases with different rhythm distributions and imbalance ratios remains an open question. Ventricular fibrillation (VF) and ventricular tachycardia (VT) are distinct arrhythmias with different ECG characteristics. VF is characterized by chaotic, high-frequency electrical activity, whereas VT is a rapid but more organized broad-complex rhythm. Despite these differences, many studies report only combined performance across both arrhythmias, making it difficult to determine whether a single feature set or imbalance-handling method performs equally well for each rhythm type. Furthermore, VT is often considered more challenging to detect than VF [24, 25], yet rhythm-specific performance is rarely reported. As a result, it remains unclear whether methods that perform well for VF generalize equally well to VT across datasets with different characteristics.

This study focuses on class imbalance as the primary object of investigation rather than treating it only as a technical challenge. Three publicly available ECG databases were combined [24, 26, 27] to increase rhythm diversity and evaluate model generalizability across datasets. To ensure a clinically meaningful and unbiased comparison, the operating point was defined before model training, and performance was assessed using recall at 95% specificity. A range of imbalance-handling approaches was compared, from a simple baseline to increasingly sophisticated methods, including inverse-frequency reweighting, LDAM loss [17], cost-sensitive learning [15], two-stage cascades, and anomaly or one-class classification [28]. All methods were evaluated under the same operating conditions using metrics that are relevant to rare-event detection, including the area under the precision–recall curve (AUPRC), missed lethal-event rate, and net clinical benefit estimated via decision curve analysis [29]. This framework enables a fair comparison of competing imbalance-handling strategies and addresses the lack of standardized evaluations in prior studies. Finally, a database-specific analysis was performed to examine differences between ventricular fibrillation (VF) and ventricular tachycardia (VT). The results show that VF is generally easier to separate from normal rhythms, whereas VT remains substantially more difficult to distinguish. These findings suggest a practical limit to the discriminative power of traditional handcrafted features and highlight the potential value of raw-waveform-based approaches for improved VT detection.

## 2 Methods

### 2.1 Datasets

Three publicly available ECG databases distributed through PhysioNet were combined for this study [30]. The VTaC dataset contains 5,037 VT alarm records; after removing segments with more than 20% missing samples, 5,013 segments were collected from ICU bedside monitors across multiple U.S. hospitals. Each 6-minute record was independently reviewed by at least two experts and classified as either a true or false VT alarm [24]. Adjudicated true VT alarms were treated as the lethal-positive class, whereas false alarms were treated as the majority class. The MIT-BIH Malignant Ventricular Ectopy Database (vfdb) consists of 22 half-hour, two-channel ECG recordings with rhythm annotations [26, 31]. Segments annotated as ventricular fibrillation (VF or VFIB), ventricular flutter (VFL), or ventricular tachycardia (VT) episodes were labelled as lethal. The Creighton University Ventricular Tachyarrhythmia Database (cudb) contains 35 eight-minute recordings with annotated VF onset and offset markers, and segments occurring within these intervals were also labelled as lethal [27]. All ECG signals sampled at 250 Hz were divided into non-overlapping 6-second windows, corresponding to 1,500 samples per segment. For continuous recordings, a segment was labelled as lethal when more than 50% of its samples occurred within an annotated VT or VF episode. After pooling all datasets, the final cohort consisted of 15,614 ECG segments, including 3,424 VT/VF segments (21.9%) and 12,190 non-lethal segments (78.1%), resulting in a normal-to-lethal ratio of approximately 3.6:1. Under this distribution, a classifier that labels every segment as normal would achieve 78.1% accuracy while failing to detect any lethal arrhythmia events.

### 2.2 Preprocessing

For each record, Lead II was selected when available, and otherwise the lead with the fewest missing samples was selected, since lead ordering is not consistent across datasets. Short missing-data gaps were filled using linear interpolation; segments exceeding this threshold were removed prior to feature extraction. Each ECG segment was then band-pass filtered (0.5–40 Hz, fourth-order zero-phase Butterworth) to remove baseline wander and high-frequency noise. After filtering, the signals were z-score normalised to standardize amplitude across recordings. Finally, each segment was cropped or zero-padded to a fixed length of 1,500 samples to ensure uniform input size for modelling.

### 2.3 Feature Extraction

Twenty-two features were computed per ECG segment to capture differences between normal rhythm, the chaotic high-frequency activity of ventricular fibrillation (VF), and the fast but relatively organized pattern of ventricular tachycardia (VT). These features were designed to summarize both temporal and spectral properties of the signal. Time-domain features were extracted directly from the ECG waveform and included mean, standard deviation, mean absolute amplitude, peak-to-peak range, robust (5th–95th percentile) range, mean absolute first and second differences, standard deviation of the first difference, zero-crossing rate, root-mean-square amplitude, skewness, kurtosis, lag-1 autocorrelation, and Hjorth mobility [32]. These measures describe signal level, variability, rate of change, regularity, and complexity of the ECG over time. Frequency-domain features were computed using Welch power spectral density [33]. These included relative band power in the 0.5–4, 4–8, 8–15, 15–30, and 30–40 Hz ranges, dominant frequency, spectral entropy, and a VF-band power ratio (4–9 Hz). These features describe how the signal energy is distributed across different frequency ranges and how structured or random the frequency content is. All features were standardized using statistics computed only from the training data to avoid information leakage. The same scaling parameters were then applied to both training and test sets.

### 2.4 Operating point and evaluation metrics

The operating point was fixed before training any model by defining performance at recall (sensitivity) at 95% specificity, corresponding to a 5% false-alarm rate consistent with typical ICU tolerances. For each model, the probability threshold achieving 95% specificity was selected from the ROC curve, and the corresponding recall was reported. Model performance was primarily summarized using AUPRC, which is more appropriate under class imbalance [5, 6], while AUROC was also reported for comparison with prior studies. The clinically most important measure was the missed-lethal-event rate (MLER = 1 recall) at the fixed operating point. Decision curve analysis was also performed to estimate net clinical benefit across a range of threshold probabilities, comparing each model against the treat-all and treat-none strategies [29]. In addition, per-class sensitivity, specificity, positive predictive value, and the full confusion matrix were reported to provide detailed performance breakdown. The dataset was split into 70% training and 30% test sets with stratification (10,929 train / 4,685 test; 1,027 lethal events in the test set), using a fixed random seed to ensure reproducibility.

### 2.5 Methods ladder

Six models were compared with increasing emphasis on the minority class (VT/VF), ranging from a simple baseline to more specialized imbalance-aware approaches.

1. Naive baseline: L2-regularised logistic regression without class weighting, used as a reference for imbalance collapse. This model treats all classes equally and does not account for rarity of VT/VF events.
2. Inverse-frequency reweighting: logistic regression with class weights inversely proportional to class frequency, assigning higher importance to VT/VF and lower weight to normal rhythms.
3. LDAM loss: a multilayer perceptron with two hidden layers (128 and 64 units), batch normalisation, ReLU activation, and dropout (0.3/0.2), trained using label-distribution-aware margin loss [17]. This enforces a larger decision margin for the minority class (max margin 0.5, scale 30). Optimization used Adam (learning rate 1e-3, weight decay 1e-4) for 30 epochs with batch size 64.
4. Cost-sensitive training: the same neural network trained with class-weighted cross-entropy, where missing a lethal event is penalised ten times more than a false alarm [15].
5. Two-stage cascade: a random forest screening model with high recall (minority-class weight 20, threshold set at the 10th percentile of positive scores to retain 90% of true events), followed by a balanced random forest applied to the selected segments to reduce false alarms.
6. Anomaly/one-class model: an Isolation Forest trained only on normal segments, where VT/VF events are detected as anomalies [28].

All models were implemented using scikit-learn [34] and PyTorch [35].

### 2.6 Statistical Analysis

Univariate feature discrimination between classes was summarized using Cohen’s d, which measures how well each individual feature separates normal and VT/VF classes by quantifying the difference between the two distributions. Per-source performance was evaluated on the held-out test set of each database separately to assess cross-database generalisation, where each dataset was tested independently to check how well the model transfers across different hospitals and data sources.

## 3 Results

### 3.1 Imbalance Problem and the Feature Space

Representative ECG waveforms (Figure 1) highlight the differences between ventricular fibrillation (VF) and ventricular tachycardia (VT). VF segments from the vfdb and cudb datasets show chaotic, high-frequency electrical activity with little organized structure, making them visually distinct from normal rhythm. In contrast, VT segments from the VTaC dataset retain a fast but organized broad-complex morphology that can resemble rapid sinus rhythm, making them more difficult to distinguish.

**Fig. 1.**
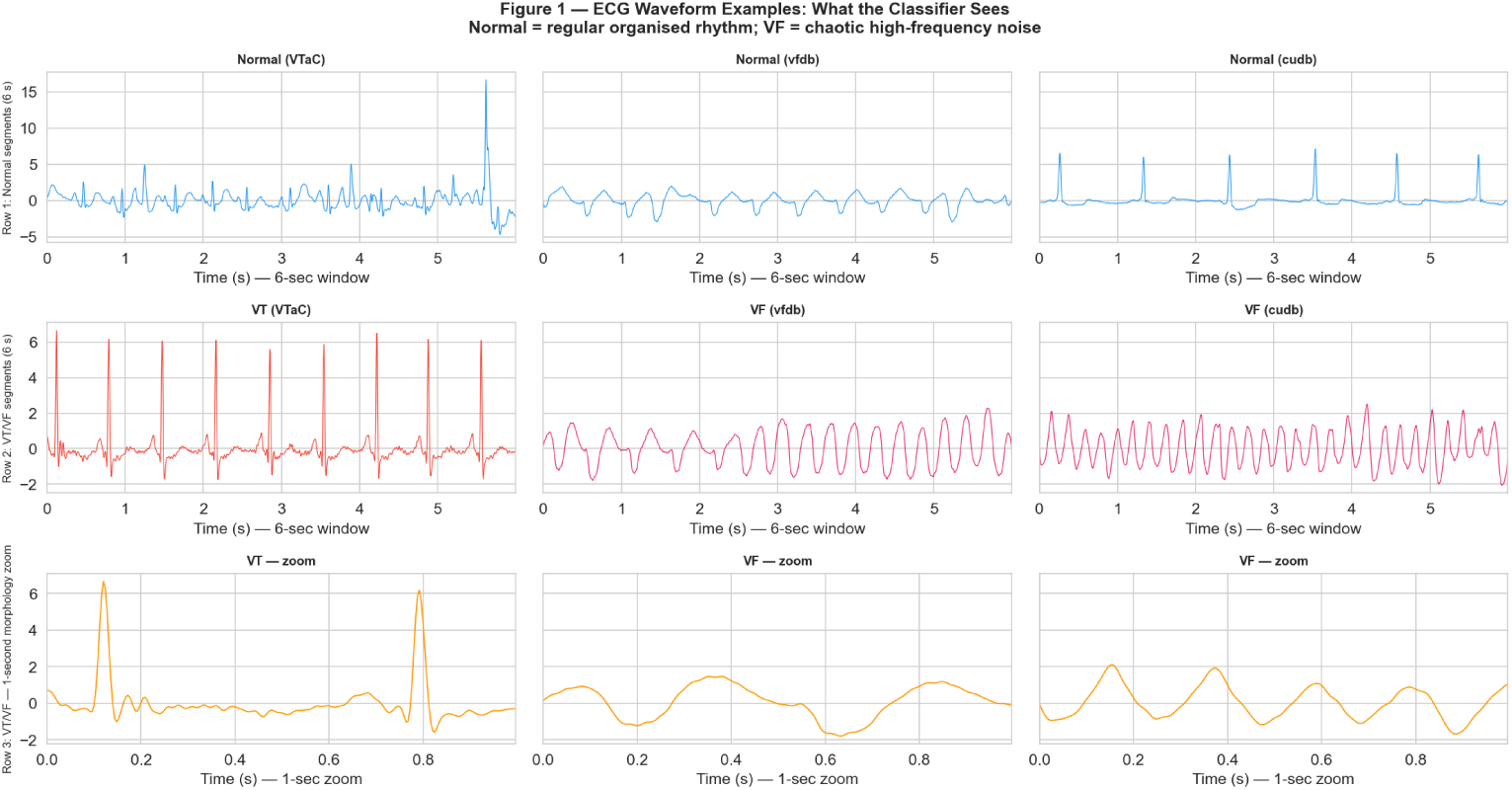
Representative ECG waveforms comparing ventricular fibrillation (VF) from the vfdb and cudb datasets with ventricular tachycardia (VT) from the VTaC dataset. VF shows chaotic, high-frequency activity, whereas VT retains an organized broad-complex morphology.

Class distribution analysis (Figure 2, Table 1) confirmed the presence of class imbalance, with an overall normal-to-lethal ratio of 3.6:1 and up to 4.6:1 in the vfdb dataset. Under these conditions, a classifier that predicts every segment as normal would still achieve 71–82% accuracy across individual datasets while failing to detect any lethal events. These findings demonstrate that accuracy alone is not an appropriate metric for evaluating models for VT/VF detection.

**Fig. 2.**
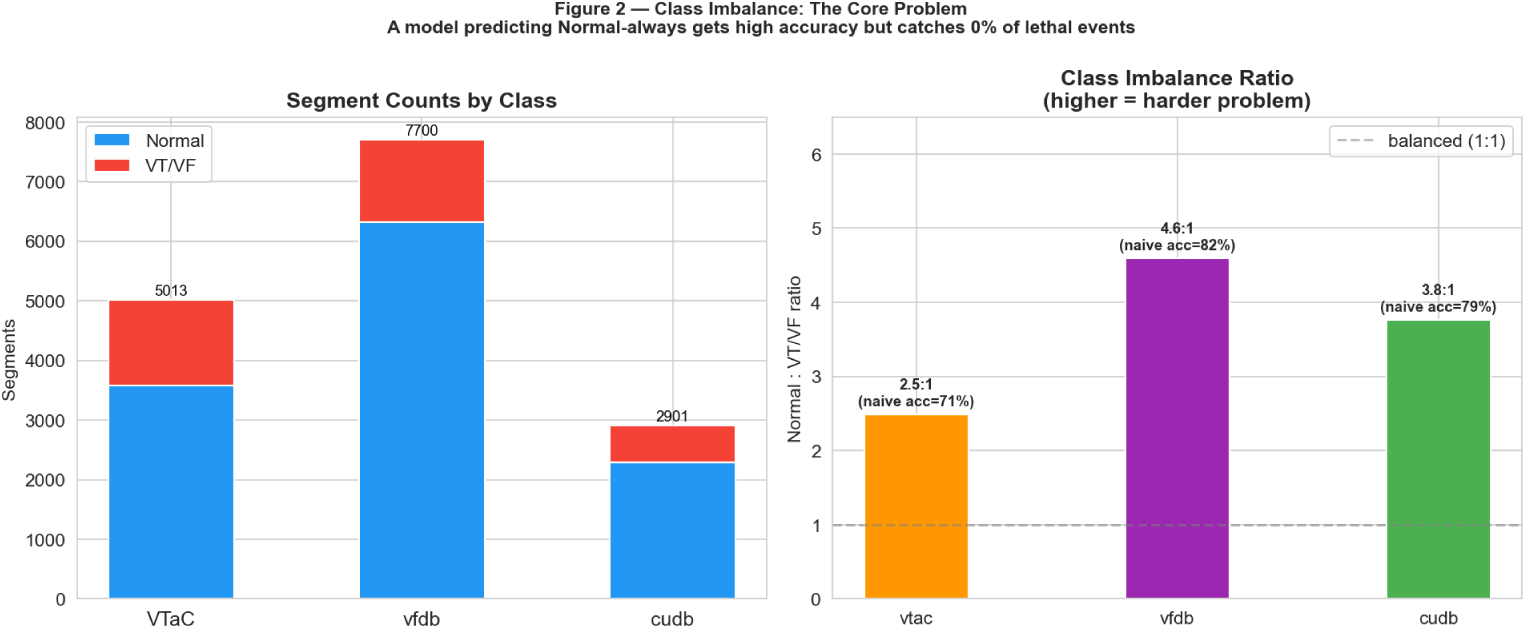
Class distribution across datasets, showing the normal-to-lethal imbalance ratio for each database.

**Table 1.** Class distribution across the three ECG databases, showing the normal-to-lethal imbalance ratio and the naive all-Normal accuracy baseline.

| Source | Total | VT/VF | Normal | Neg:Pos | VT/VF % | Naive accuracy |
| --- | --- | --- | --- | --- | --- | --- |
| vtac | 5013 | 1438 | 3575 | 2.5:1 | 28.7% | 71.3% |
| vfdb | 7700 | 1377 | 6323 | 4.6:1 | 17.9% | 82.1% |
| cudb | 2901 | 609 | 2292 | 3.8:1 | 21.0% | 79.0% |
| ALL | 15614 | 3424 | 12190 | 3.6:1 | 21.9% | 78.1% |

Features were ranked by effect size (Figure 3, Table 2) using Cohen’s *d* to measure how well each individual feature separated normal rhythm from VT/VF. The most discriminative features were spectral entropy (*d* = 0.52), mean absolute amplitude (0.49), dominant frequency (0.46), and 8–15 Hz band power (0.44). These features mainly capture the chaotic, high-frequency electrical activity that is characteristic of ventricular fibrillation (VF). However, none of the features achieved more than a medium effect size. The highest Cohen’s *d* value of 0.52 indicates that, although the handcrafted features are physiologically meaningful, each feature alone provides only moderate separation between normal and VT/VF rhythms. This suggests that the feature set has an inherent limitation, and even more advanced classifiers may not substantially improve recall, particularly for the more challenging VT cases.

**Fig. 3.**
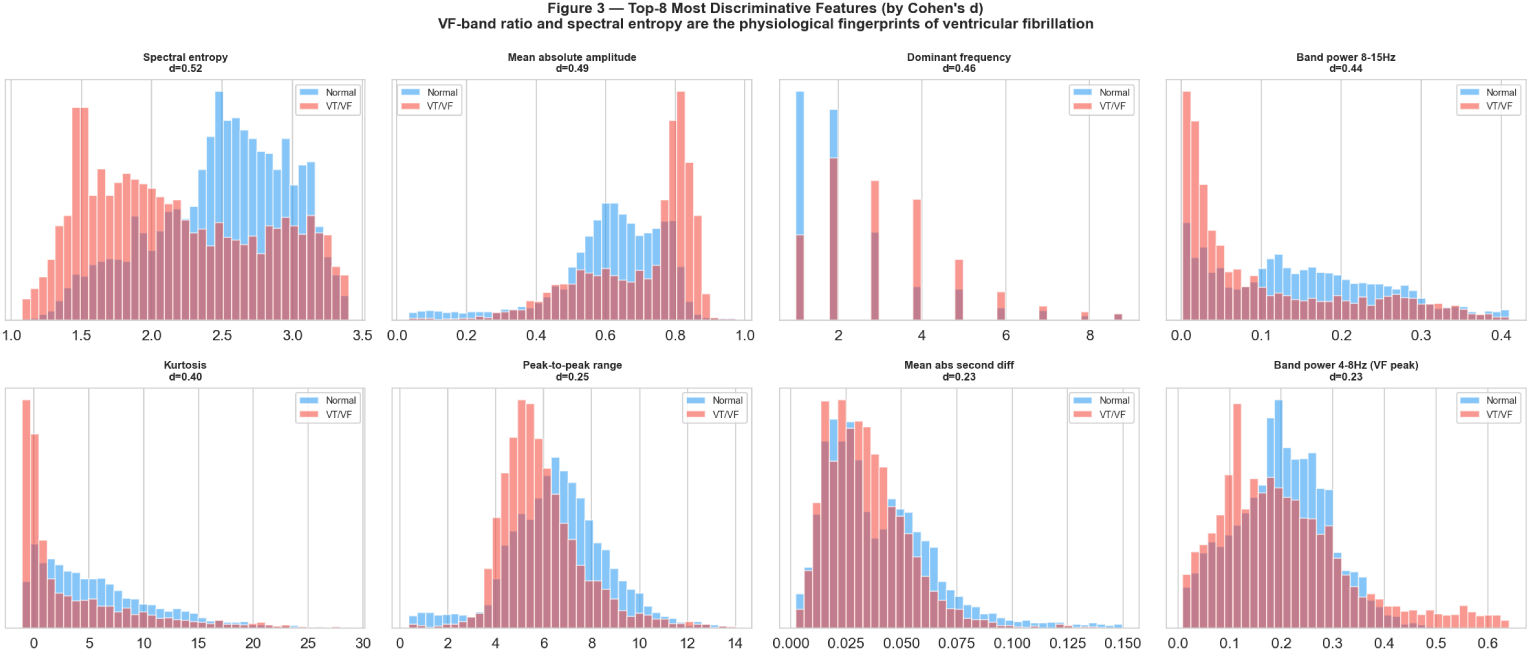
Distributions of the most discriminative handcrafted features for normal versus VT/VF rhythms, ranked by Cohen’s *d* effect size.

**Table 2.** Handcrafted features ranked by Cohen’s *d* effect size for separating normal rhythm from VT/VF.

| Feature | Cohen’s $d$ |
| --- | --- |
| Spectral entropy | 0.519 |
| Mean absolute amplitude | 0.486 |
| Dominant frequency | 0.461 |
| Band power 8–15 Hz | 0.444 |
| Kurtosis | 0.398 |
| Peak-to-peak range | 0.249 |
| Mean abs second diff | 0.231 |
| Band power 4–8 Hz (VF peak) | 0.226 |
| Band power 0.5–4 Hz | 0.201 |
| Hjorth mobility | 0.184 |
| Robust range (P5–P95) | 0.177 |
| Std of diff | 0.176 |
| VF-band ratio (4–9 Hz) | 0.151 |
| Band power 15–30 Hz | 0.143 |
| Band power 30–40 Hz | 0.136 |
| Zero-crossing rate | 0.126 |
| RMS | 0.124 |
| Std amplitude | 0.124 |
| Lag-1 autocorrelation | 0.123 |
| Skewness | 0.100 |
| Mean abs first diff | 0.043 |
| Mean amplitude | 0.003 |

### 3.2 Naive Collapse and the Methods Ladder

Before evaluating the models, the operating point was fixed at 95% specificity, allowing only 5% of normal rhythms to be incorrectly classified as VT/VF. At this operating point, the recall of each model was measured to determine how many lethal events were correctly detected. Models were then compared with increasing emphasis on the minority class to assess whether imbalance-aware approaches could reduce the number of missed VT/VF events.

The naive logistic regression model was first evaluated without any imbalance handling. Although it achieved an AUROC of 0.822 and an accuracy of 84.1%, these results appeared better than the model’s actual ability to detect lethal events. At the fixed operating point of 95% specificity, only 45.1% of VT/VF events were correctly detected, while 564 of the 1,027 lethal events were missed (MLER = 0.549; Table 3, Figure 4). These findings show that high AUROC and accuracy alone do not necessarily reflect good performance for detecting clinically important events. Inverse-frequency reweighting was then applied by assigning a higher weight to the minority class during training. However, this approach produced only a minimal improvement. The AUROC increased slightly to 0.825, recall improved from 45.1% to 45.5%, and only four additional VT/VF events were detected compared with the baseline. These results suggest that, even with a moderate class imbalance of 3.6:1, simply adjusting class weights is not sufficient to improve VT/VF detection. More advanced methods that improve the separation between the two classes are therefore needed, which motivated the evaluation of the remaining imbalance-aware models.

**Fig. 4.**
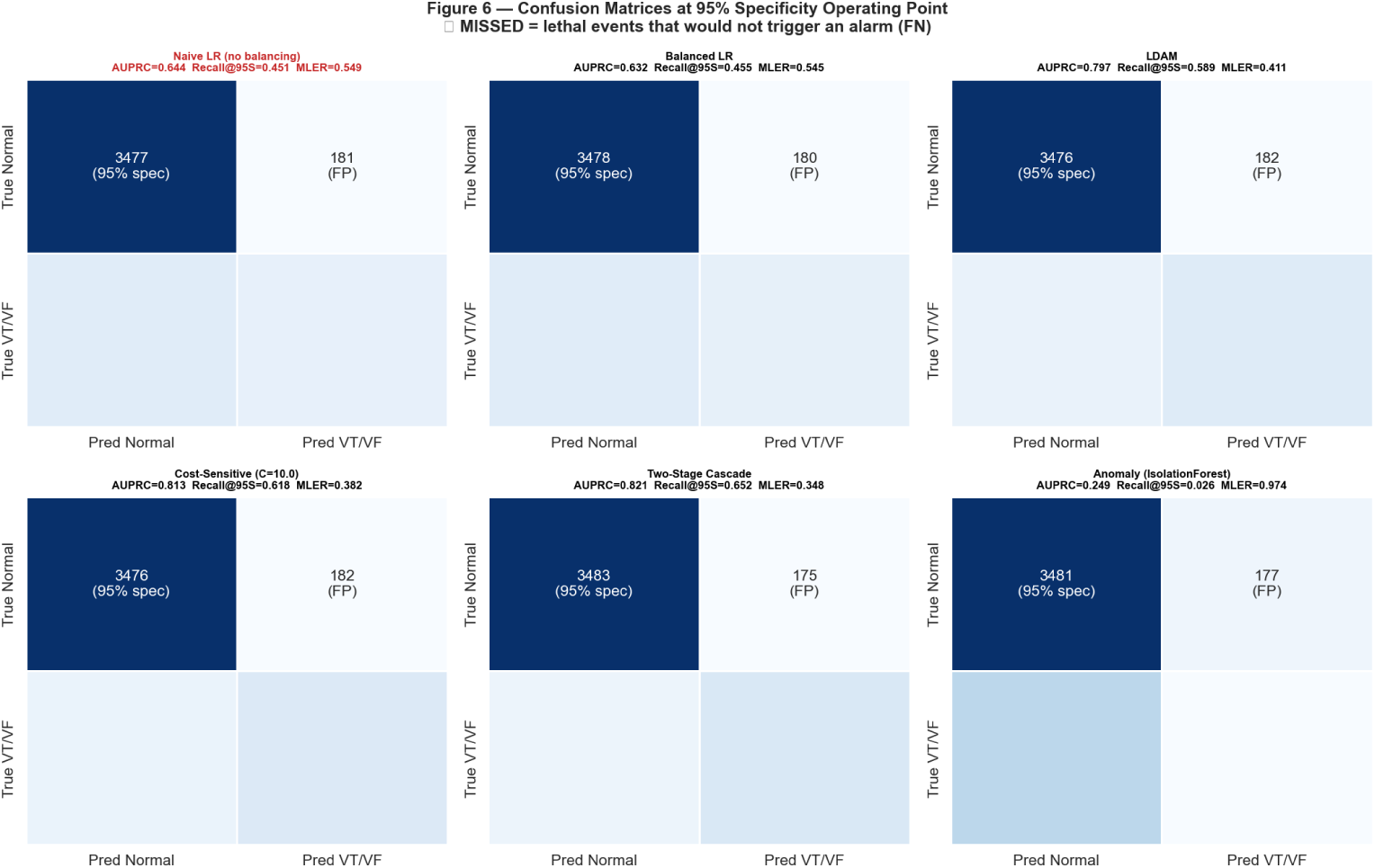
Confusion matrices at the fixed operating point of 95% specificity, showing detected versus missed VT/VF events and false alarms for each model.

**Table 3.**
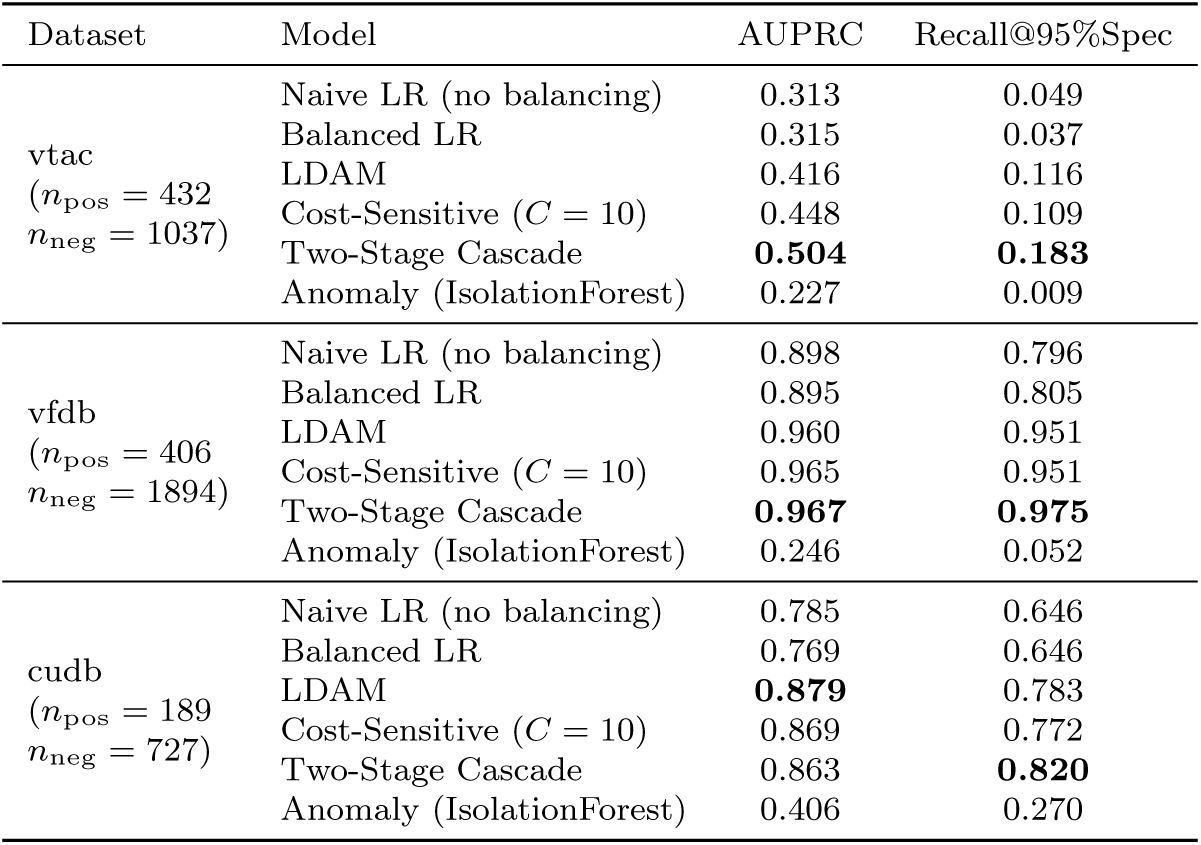
Imbalance-handling model performance across the three ECG databases, evaluated by AUPRC and recall at 95% specificity. The best value per dataset for each metric is shown in bold. *n*_pos_ and *n*_neg_ denote the number of positive (VT/VF) and negative (Normal) test segments.

Methods that modified the learning objective or model architecture showed a clear improvement in recall compared with the simpler approaches (Table 4, Figure 5, Figure 6, Figure 7). LDAM increased recall to 58.9% (MLER = 0.411, AUPRC = 0.797), while cost-sensitive training further improved recall to 61.8% (MLER = 0.382, AUPRC = 0.813) and achieved the highest AUROC (0.927). These results indicate that imbalance-aware training methods improve detection of VT/VF events. Among all models, the two-stage cascade achieved the best performance at the fixed operating point of 95% specificity. It reached an AUPRC of 0.821, a recall of 65.2%, and an MLER of 0.348. Compared with the naive baseline, the number of missed lethal events decreased from 564 to 357, representing a 37% reduction, while maintaining a similar number of false alarms (175 vs. 181; Figure 4). As expected, the anomaly/one-class model performed poorly, detecting only 2.6% of VT/VF events (MLER = 0.974). This result suggests that training with labeled VT/VF examples is necessary for effective detection. The Precision–Recall curves (Figure 6) distinguished the models more clearly than the ROC curves under class imbalance. The two-stage cascade and cost-sensitive models maintained higher precision while achieving substantially higher recall than the other approaches.

**Fig. 5.**
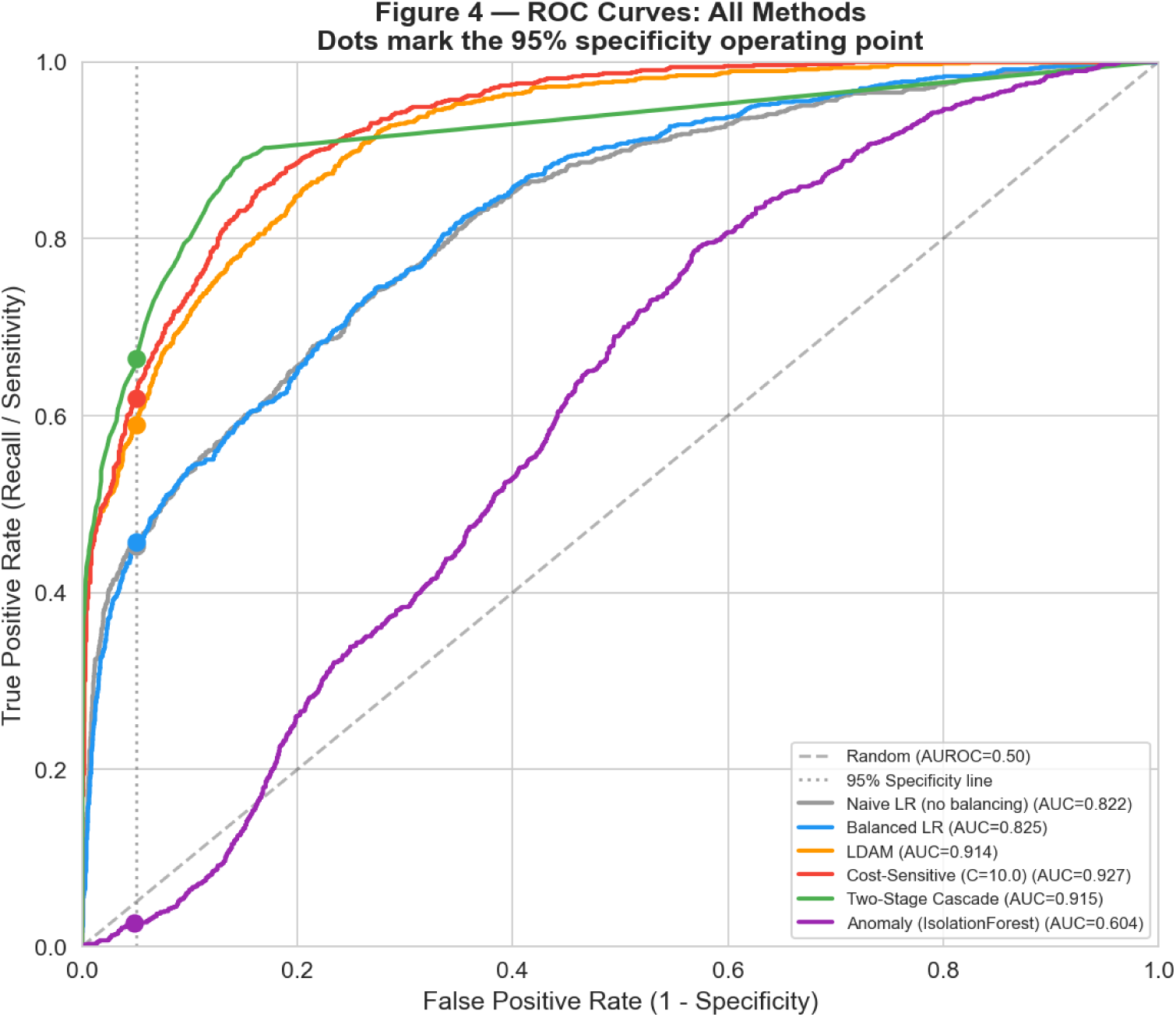
Receiver operating characteristic (ROC) curves for the evaluated models on the pooled dataset.

**Fig. 6.**
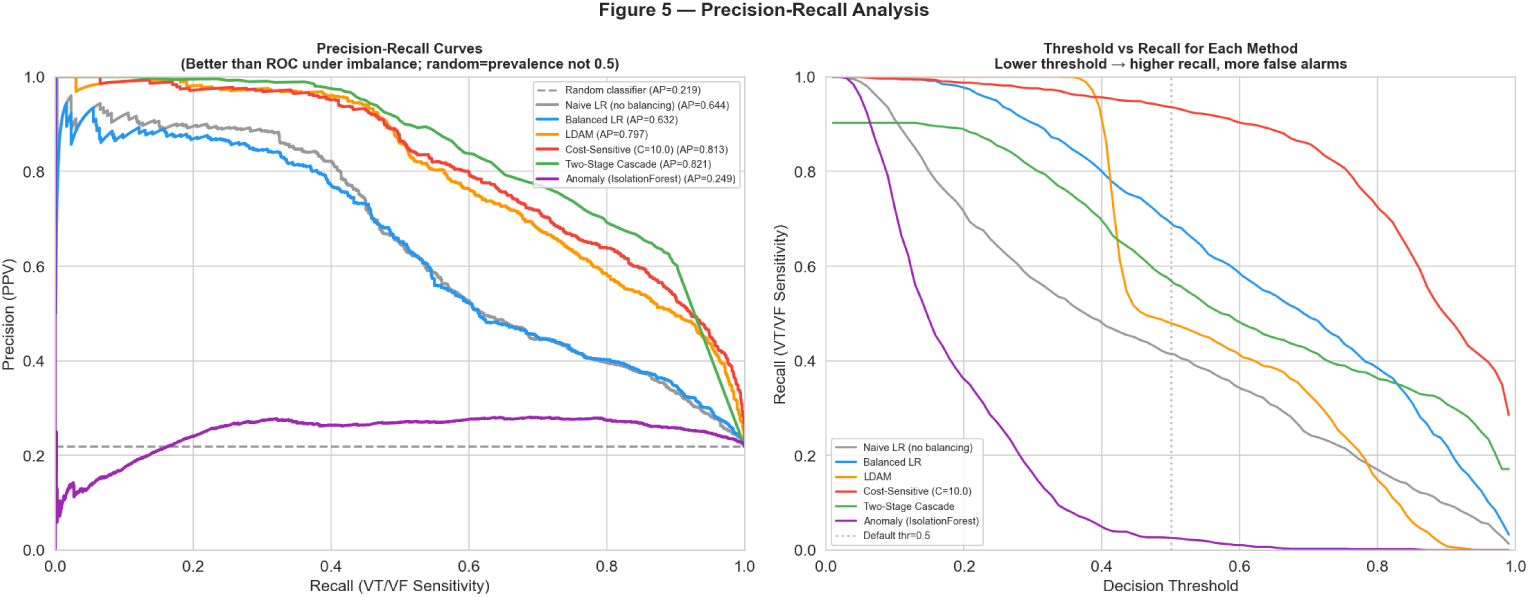
Precision–Recall (PR) curves for the evaluated models. Under class imbalance, the PR curves separate model performance more clearly than the ROC curves.

**Fig. 7.**
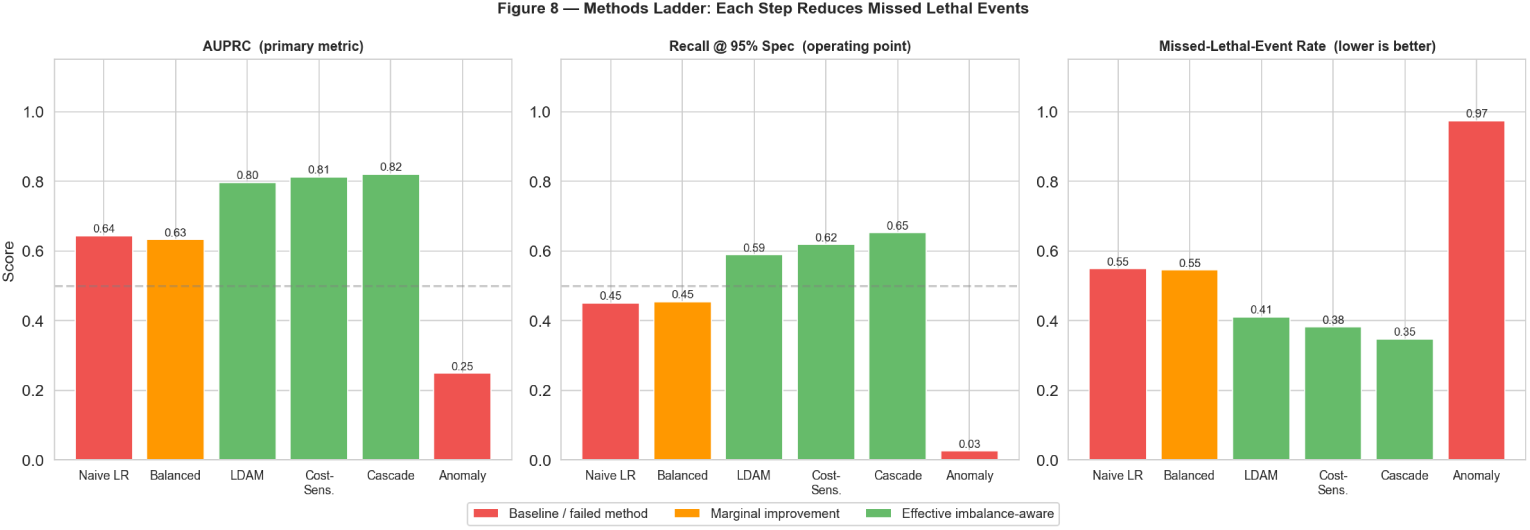
Imbalance-handling methods arranged as a ladder of increasing emphasis on the minority class, from the naive baseline through inverse-frequency reweighting, LDAM, cost-sensitive learning, and the two-stage cascade, showing the corresponding improvement in recall at 95% specificity.

**Table 4.** Pooled classification performance at the fixed operating point of 95% specificity. AUROC and AUPRC summarise overall discrimination; sensitivity equals recall for the VT/VF class; MLER is the missed lethal-event rate; Missed (FN) is the number of missed VT/VF events out of 1,027 total positives. The best value per metric is shown in bold.

| Model | AUROC | AUPRC | Sensitivity | Specificity | MLER | Missed |
| --- | --- | --- | --- | --- | --- | --- |
| Naive LR (no balancing) | 0.822 | 0.644 | 0.451 | 0.951 | 0.549 | 564 |
| Balanced LR | 0.825 | 0.632 | 0.455 | 0.951 | 0.545 | 560 |
| LDAM | 0.914 | 0.797 | 0.589 | 0.950 | 0.411 | 422 |
| Cost-Sensitive ($C = 10$) | 0.927 | 0.813 | 0.618 | 0.950 | 0.382 | 392 |
| Two-Stage Cascade | 0.915 | **0.821** | **0.652** | 0.952 | **0.348** | **357** |
| Anomaly (IsolationForest) | 0.604 | 0.249 | 0.026 | 0.952 | 0.974 | 1000 |

### 3.3 Per-Source Generalisation and Clinical Net Benefit

Performance was decomposed by source (Figure 8, Table 3), revealing a clear and physiologically consistent difference between datasets. On VF-dominated databases, the cascade model detected most events, achieving a recall of 0.975 on vfdb and 0.820 on cudb (AUPRC 0.967 and 0.863, respectively). In contrast, on the VT-alarm database (VTaC), performance was substantially lower, with a recall of 0.183 (AUPRC 0.504). This pattern was consistent across all evaluated methods. The difference is driven by the underlying signal characteristics rather than the choice of algorithm. Ventricular fibrillation (VF) exhibits chaotic, high-frequency activity that is well captured by spectral entropy, dominant frequency, and band-power features, making it highly separable from normal rhythm. In contrast, ventricular tachycardia (VT) is a fast but organized broad-complex rhythm whose spectral properties overlap with rapid sinus rhythm, limiting separation using hand-crafted features. As a result, the overall recall ceiling of approximately 65% is primarily determined by the difficulty of detecting VT events.

**Fig. 8.**
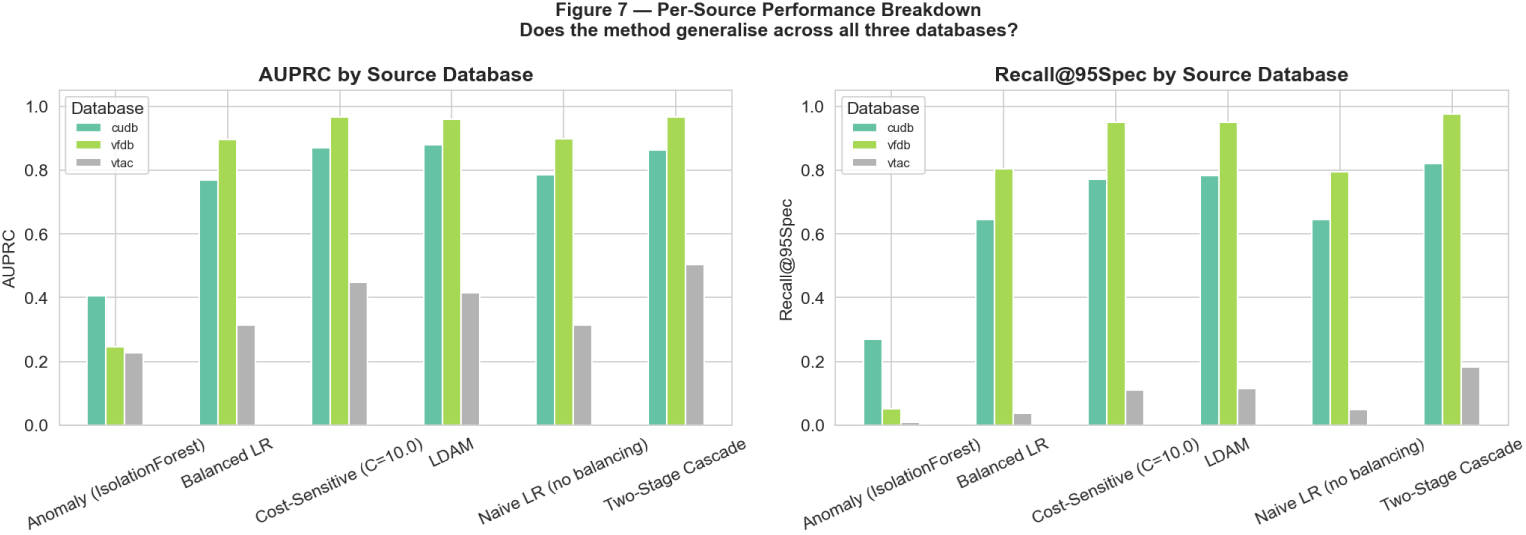
Per-database model performance, showing AUPRC and recall at 95% specificity for each imbalance-handling method across the vtac, vfdb, and cudb datasets. VF-dominated databases (vfdb, cudb) are more separable than the VT-dominated vtac database across all methods.

Decision-curve analysis (Figure 9) was used to translate model performance into clinical usefulness, assessing whether the models provide net benefit in practice rather than only statistical accuracy. Across most plausible alarm-threshold settings, the two-stage cascade model achieved the highest net benefit compared with all other methods. It remained consistently above both reference strategies, namely “alarm-on-everything” and “never-alarm,” which represent the extremes of excessive and absent alerting. In contrast, some other models dropped below these reference strategies at certain thresholds, indicating limited clinical utility in those settings. Cost-sensitive training showed competitive performance at lower thresholds where more aggressive alarming is allowed; however, the two-stage cascade remained the most stable and consistently beneficial approach across operating conditions. Standard performance metrics, including sensitivity, specificity, and positive predictive value at the chosen operating point, are reported in Table 4.

**Fig. 9.**
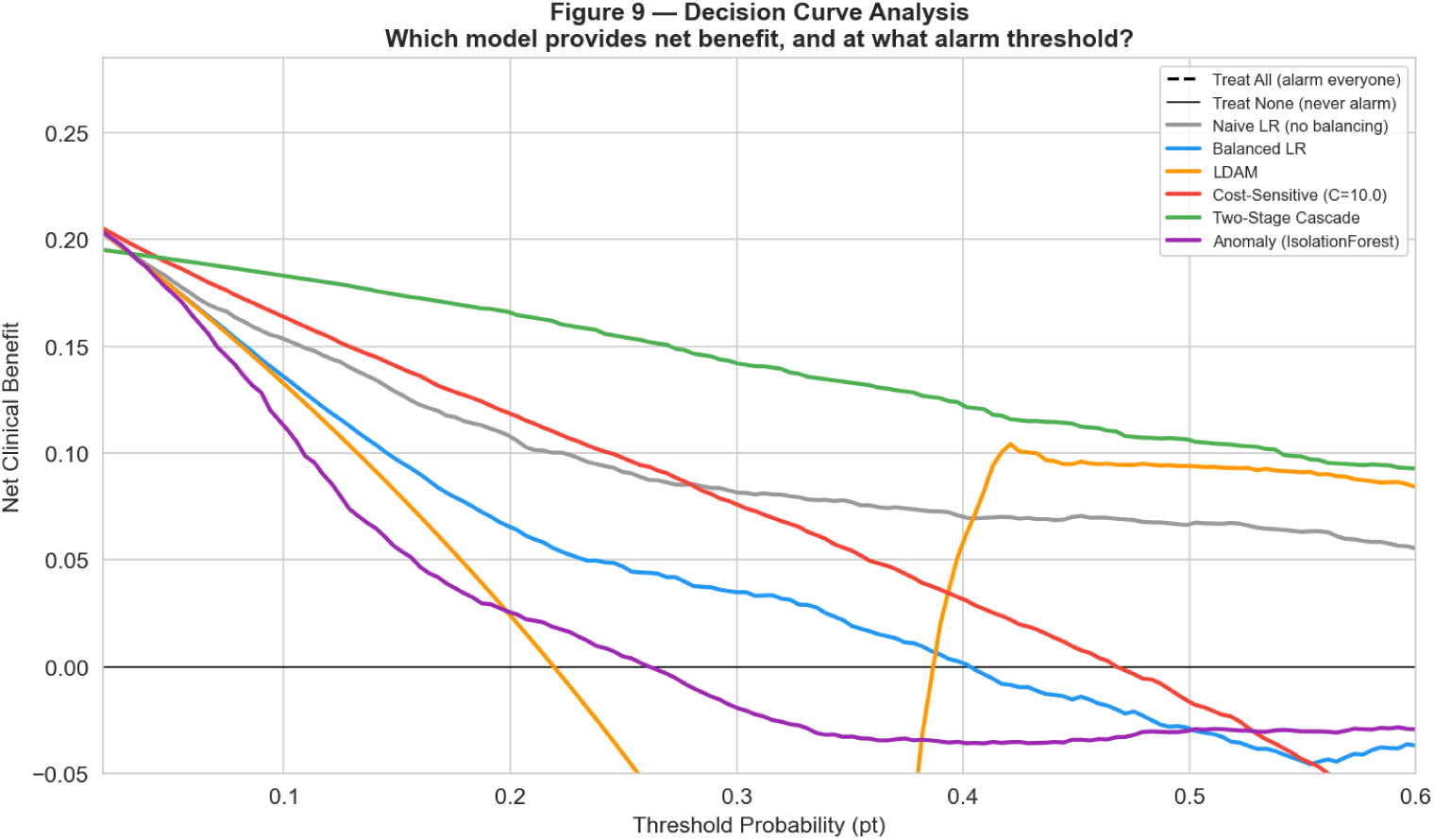
Decision curve analysis showing net clinical benefit across a range of threshold probabilities for the evaluated models.

## 4 Discussion

Across three public ECG databases, it was shown that accuracy and AUROC are not reliable success criteria for lethal-arrhythmia detection. A baseline model that appeared acceptable under these metrics still missed more than half of all lethal events when evaluated at a realistic specificity level. This indicates that high accuracy and AUROC do not necessarily reflect safe clinical performance. The most common baseline correction, inverse-frequency reweighting, showed almost no improvement under moderate class imbalance. In contrast, methods that modified either the learning objective (LDAM, cost-sensitive training) or the model structure (two-stage cascade) produced consistent reductions in missed events. Among all approaches, the two-stage cascade achieved the best performance, reducing missed lethal events by approximately 37% and providing the highest decision-curve net benefit. The remaining errors were concentrated in a specific physiologically interpretable limitation, suggesting that the failure mode is driven by signal characteristics rather than model design.

The remaining errors were mainly caused by the characteristics of the ECG signals rather than the machine learning models. Ventricular fibrillation (VF) has a chaotic, high-frequency electrical pattern, which is well captured by features such as spectral entropy, dominant frequency, and the 4–9 Hz VF-band power ratio. As a result, VF was detected with a recall of up to 0.975. In contrast, ventricular tachycardia (VT) is a fast but organized broad-complex rhythm. Its frequency characteristics overlap with those of rapid sinus rhythm, making it difficult for the same handcrafted features to distinguish the two. Consequently, recall on the VT database was only 0.183, indicating that the handcrafted feature set is not sufficient for reliable VT detection. These findings are consistent with previous studies showing that VT is one of the most difficult life-threatening arrhythmias to detect reliably [24]. They also agree with earlier work reporting that morphology- and template-based features provide only limited separation of VT from other rhythms [25].

These findings provide practical guidance for the design and evaluation of lethal arrhythmia detection systems. Such systems should be evaluated at a fixed clinical operating point and report the missed-lethal-event rate rather than relying mainly on accuracy, because a model with high accuracy can still miss many dangerous events [1]. When labelled VT/VF data are available, cost-sensitive training and two-stage cascade models should be preferred over simple inverse-frequency reweighting, as they provide better detection of lethal events. Model selection should also consider clinical net benefit instead of relying only on discrimination metrics such as accuracy or AUROC [29]. Performance should also be evaluated separately for different arrhythmia types. Spectral feature-based models detect ventricular fibrillation (VF) reliably but perform much less effectively for organised ventricular tachycardia (VT). This difference should be considered when configuring alarm systems and highlights the need for improved methods to detect VT. Previous studies have reported good overall performance using different methods to handle class imbalance, including oversampling, focal loss, CNN-BLSTM models, and ensemble methods [13, 16, 19–22]. High accuracy has also been reported for dedicated shockable-rhythm detectors, particularly for ventricular fibrillation (VF), which is generally easier to distinguish from normal rhythm [23]. The findings of this study are consistent with previous work for VF but extend them in two important ways. First, all models were evaluated at the same fixed operating point (95% specificity), and performance was assessed using the missed-lethal-event rate instead of relying mainly on accuracy or F1 score. This showed that different imbalance-handling methods can produce large differences in the number of lethal events detected, even when their overall performance metrics appear similar. Second, performance was evaluated separately for each arrhythmia type. This showed that the main limitation in detecting organised ventricular tachycardia (VT) is the handcrafted feature representation rather than the imbalance-handling method. These findings suggest that improving class imbalance alone is unlikely to substantially improve VT detection, and more informative features or advanced models may be needed.

This study has several limitations. First, although the best model achieved an overall recall of 65%, this performance is not sufficient for clinical use. The study should therefore be considered a methodological proof of concept rather than a deployable clinical system. Second, the main limitation lies in the feature representation rather than the machine learning models. The highest univariate effect size was only 0.52, indicating that the 22 handcrafted features cannot clearly distinguish organised ventricular tachycardia (VT) from rapid sinus rhythm. Third, the evaluation was performed on individual ECG segments using a single lead rather than complete patient recordings or multi-lead ECG signals. Future studies should investigate whether patient-level evaluation and additional ECG leads improve performance. Fourth, the three databases used different definitions of the positive class. For example, the VTaC dataset defines positive cases as expert-confirmed true VT alarms rather than a direct comparison between VT and sinus rhythm. This difference should be considered when interpreting the results across datasets. Finally, the study was performed retrospectively using public databases. Additional validation on independent datasets and prospective continuous ECG monitoring is needed before these methods can be considered for clinical application.

The next step is to use deep learning models that learn directly from raw ECG waveforms instead of relying on handcrafted features. One-dimensional convolutional neural networks and attention-based models can learn waveform patterns that handcrafted spectral features may miss, particularly for VT [9, 10, 12, 36]. Future studies should also continue using the evaluation framework established in this study by assessing models at a fixed operating point and reporting the missed-lethal-event rate. In addition, incorporating multi-lead ECG signals, evaluating complete patient recordings, and validating the models prospectively on continuous monitoring data could help move this proof-of-concept toward a clinically deployable detector.

## 5 Conclusion

Detecting rare lethal arrhythmias is fundamentally a class-imbalance problem, and the choice of evaluation metrics determines whether this issue is visible or hidden. In this study across three public ECG databases, accuracy and AUROC appeared acceptable for a naive classifier but masked a missed-lethal-event rate above 50%. A simple inverse-frequency reweighting approach did not substantially improve this limitation. In contrast, imbalance-aware methods that modify model structure performed better, with the two-stage cascade achieving the strongest results by reducing missed events by approximately 37% while improving clinical net benefit at a fixed operating point.

The residual error is mainly concentrated in organized ventricular tachycardia, which has strong spectral similarity to fast sinus rhythm and therefore creates a feature-space ceiling. This means most remaining misclassifications occur because VT and fast normal rhythms look too similar in the feature representation, limiting how well any model can separate them. For clinicians and patients relying on continuous ECG monitoring, the key implication is that performance of lethal-arrhythmia detectors should be evaluated primarily by the number of missed events rather than overall correctness. Improving detection of VT will likely require models that learn directly from raw ECG waveforms instead of relying only on handcrafted features.

## Data Availability

All data used in this study are publicly available from PhysioNet. The study used the VTaC (Ventricular Tachycardia Alarms in the ICU), MIT-BIH Malignant Ventricular Ectopy Database (VFDB), and Creighton University Ventricular Tachyarrhythmia Database (CUDB). No new patient data were generated. Source databases were openly available prior to the initiation of this study and can be accessed at the links provided.

https://physionet.org/content/vtac/1.0/

https://physionet.org/content/cudb/1.0.0/

